# Atlas-Based MRI Spatial Phenotyping in Pediatric Low-Grade Glioma

**DOI:** 10.64898/2026.10.29.26366718

**Authors:** Bhavyasri Vunnava, Lu Men, Fadel Batal, Dimosthenis Chrysochoou, Adam Kraya, Deep Gandhi, Phillip B. Storm, Peter J. Madsen, Adam Resnick, Arastoo Vossough, Ali Nabavizadeh, Anahita Fathi Kazerooni

## Abstract

Pediatric low-grade gliomas (pLGGs) arise throughout the developing brain, and their anatomic distribution is related to age, molecular subtype, and extent of resection (EOR). In clinical studies, pLGG location is commonly represented by a single manually assigned anatomic category, which does not quantify the full spatial extent of tumors spanning multiple brain regions. We developed and evaluated a reproducible, publicly available pediatric atlas-based framework integrating voxel-wise probabilistic mapping with patient-level atlas-derived spatial features. In this retrospective study, preoperative MRI from 349 patients with pLGG was registered to the Haskins pediatric template using EasyReg. Probabilistic maps showed differences in tumor distribution by age, molecular subtype, and EOR, including greater optic pathway and suprasellar involvement at 0–2 years and greater cerebellar involvement in *KIAA1549–BRAF* fusion tumors. Atlas-derived spatial, tumor morphometric, and clinical features distinguished gross total resection/near total resection from subtotal resection or biopsy/no-surgery (Replication area under the curve, 0.864). Substituting 107 automatically computed regional overlap features for the manually assigned location variable in previously published clinico-imaging risk stratification models did not significantly alter discrimination (smallest *P* = 0.12). Together, these group- and patient-level analyses demonstrate the framework’s utility for quantitative spatial phenotyping in imaging and clinical studies of pLGG.

## Introduction

Pediatric low-grade glioma (pLGG) is the most common central nervous system tumor in children, and these tumors arise throughout the developing brain [1,2]. Their anatomic distribution is closely related to age, molecular subtype, and extent of resection (EOR) [2–4]. EOR is in turn associated with progression-free survival (PFS) [3]. Because overall survival in pLGG is high, the anatomic factors associated with EOR and progression are of particular clinical importance [5]. In clinical studies, pLGG location is commonly represented using discrete anatomic categories [3,5]. In previously published clinico-imaging risk stratification models, tumor location is included as a single variable, assigned manually for each patient [6,7]. Although useful, these categories do not quantify the full spatial extent of tumors that span multiple brain regions.

MRI-based voxel-wise probabilistic mapping has been used in adult glioma and other intracranial tumors to examine associations between tumor location and molecular, clinical, management, and outcome characteristics [8–10]. Quantitative mapping of tumor location has also been applied in pLGG, including three-dimensional spatial probability distributions for *BRAF* molecular subtype classification [11]. However, that study addressed a single classification task and did not provide a framework for spatial phenotyping across age, molecular subtype, and EOR. Moreover, the spatial representation was derived in an adult reference space rather than a pediatric atlas. Because pediatric templates more closely reflect pediatric brain anatomy and may require less deformation during registration than adult templates [12,13], a pediatric atlas-based framework integrating voxel-wise probabilistic mapping with patient-level atlas-derived spatial features could provide a reproducible, quantitative spatial representation of these relationships rather than categorical localization.

The purpose of this study was to develop and evaluate a reproducible and publicly available pediatric atlas-based framework for spatial phenotyping of pLGG using the Haskins pediatric template [13] and EasyReg, a deep learning–based method for multimodal brain MRI registration. We used the framework to generate voxel-wise probabilistic maps by age, molecular subtype, and EOR, with additional maps by sex. We then tested whether atlas-derived spatial, tumor morphometric, and clinical features could distinguish gross total resection/near total resection (GTR) from subtotal resection (STR) or biopsy/no-surgery. Finally, we evaluated whether the manually assigned tumor location variable could be replaced with 107 atlas-derived regional overlap features in previously published clinico-imaging risk stratification models [6,7].

## Methods

### Study Sample

This retrospective, HIPAA–compliant study analyzed imaging, clinical, and molecular data obtained from the Children’s Brain Tumor Network (CBTN) [14], a publicly accessible biorepository. Patients were diagnosed between October 2001 and January 2024. Patients were included if they had pLGG at initial diagnosis and a complete set of four standard MRI sequences: T1-weighted, contrast-enhanced T1-weighted, T2-weighted, and FLAIR. These imaging inclusion criteria were retained from the previously published imaging cohort to enable evaluation of atlas-derived tumor location features within the same clinico-imaging risk stratification models [7]. Patients with substantial motion artifacts or tumors located outside the brain were excluded, resulting in an imaging cohort of 360 patients. This cohort of 360 patients, of whom 349 were analyzed, overlaps with those used in the two studies that developed these models [6,7]. However, the pediatric atlas-based framework and spatial analyses reported here were not part of either study.

### Study Overview

Figure 1 summarizes the study design. Preoperative MRI and tumor segmentations were registered to the Haskins pediatric template, and voxel-wise probabilistic maps and atlas-derived spatial features were generated in Haskins template space. Atlas-derived spatial features, together with tumor morphometric and clinical features, were used for classification of EOR, and the manually assigned tumor location variable in previously published clinico-imaging risk stratification models was replaced with the 107 atlas-derived regional overlap features [6,7].

**Figure 1.**
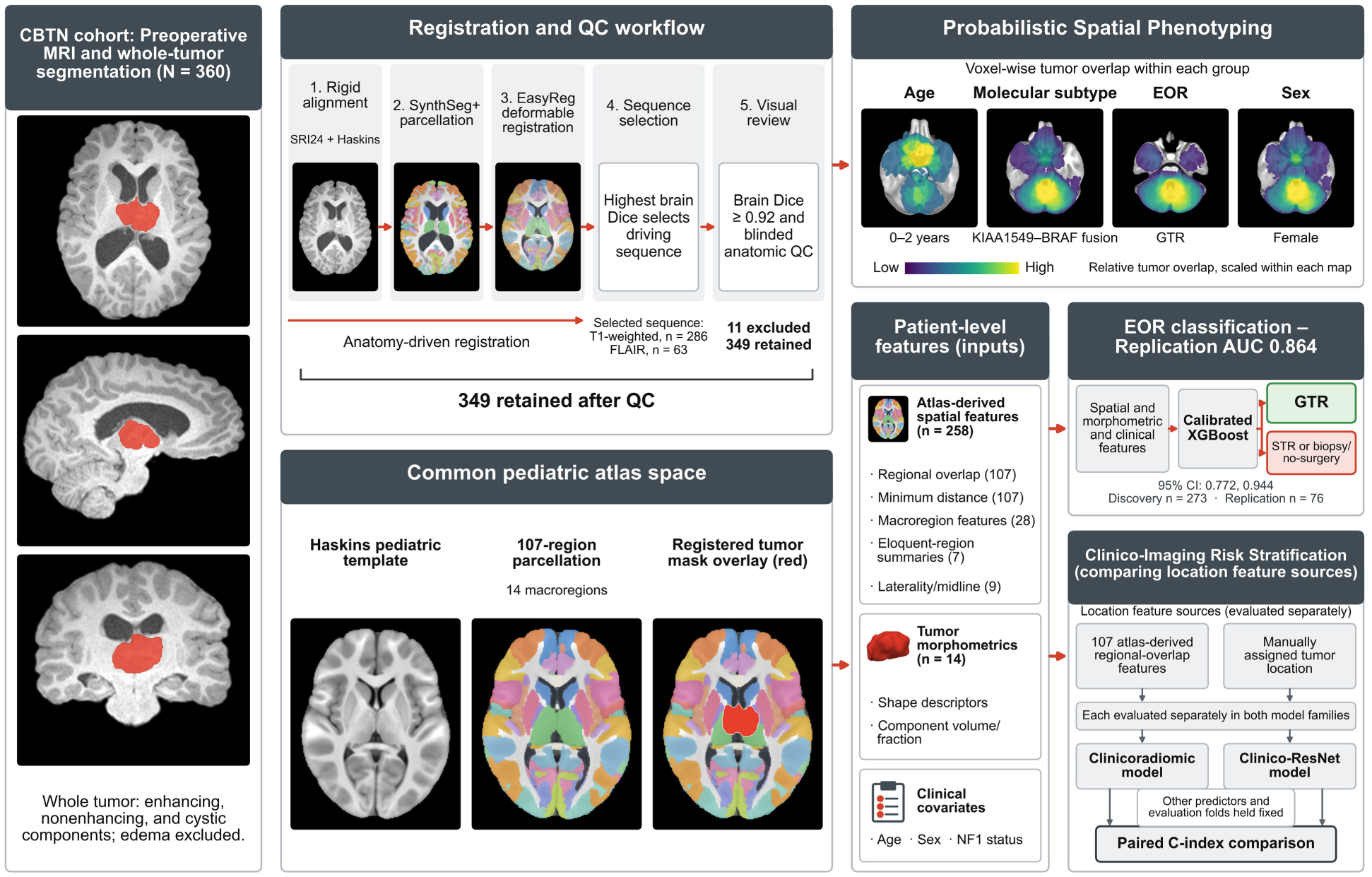
Atlas-based MRI spatial phenotyping in pediatric low-grade glioma (pLGG). Preoperative MRI and whole tumor segmentations (enhancing, nonenhancing, and cystic components, edema excluded) from 360 patients in the Children’s Brain Tumor Network (CBTN) cohort were registered to the Haskins pediatric template by rigid alignment from SRI24 space, SynthSeg+ parcellation, and EasyReg deformable registration. The sequence with the highest brain Dice was retained for each patient, and blinded visual review at a brain Dice threshold of 0.92 excluded 11 patients, leaving 349 (T1-weighted, n = 286; FLAIR, n = 63). In the common pediatric atlas space, registered tumor masks were averaged within each group to form voxel-wise probabilistic maps by age, molecular subtype, extent of resection (EOR), and sex, shown here for children aged 0–2 years, *KIAA1549–BRAF* fusion tumors, tumors with GTR, and female patients. Each tumor was also summarized as patient-level atlas-derived spatial features, tumor morphometric features, and clinical covariates. These features were used for classification of EOR (GTR versus STR or biopsy/no-surgery), and the manually assigned tumor location variable was replaced with the 107 atlas-derived regional overlap features in the clinicoradiomic and clinico-ResNet models, compared by paired concordance index. GTR = gross total resection/near total resection, STR = subtotal resection, FLAIR = fluid-attenuated inversion recovery, AUC = area under the receiver operating characteristic curve, CI = confidence interval, C-index = concordance index, NF1 = neurofibromatosis type 1, QC = quality control.

### MRI Segmentation and Clinical Variables

Brain tumor segmentations were generated using a validated pediatric deep learning segmentation method [15] and reviewed by trained readers and a board-certified neuroradiologist blinded to molecular subtype and outcome. The whole tumor mask was defined as the union of enhancing, nonenhancing, and cystic components; peritumoral edema was excluded. Clinical and molecular variables included age, sex, EOR, molecular subtype, and PFS. EOR was categorized as GTR (i.e., gross or near total resection, as provided through CBTN), STR, or biopsy/no-surgery as assigned at the institutions contributing to CBTN, and no uniform definition was applied centrally. PFS was obtained from CBTN; events included radiographic progression, recurrence, or death, with censoring at last follow-up.

### Pediatric Atlas Registration and Quality Control

Images were preprocessed as previously described [6], and the preprocessing steps are detailed in the Supplementary Methods. Because preprocessing had already placed every image in SRI24 space, a single rigid transform from SRI24 space to the Haskins pediatric template [13] was computed with FreeSurfer mri_robust_register [16] and applied to every patient. Each available structural MRI sequence and the template were parcellated with SynthSeg+ [17], and the sequence was deformably registered to the template with EasyReg [18,19] using those parcellations. Registration quality was assessed for each sequence using brain Dice, the overlap between the whole-brain masks derived from the registered patient and template parcellations (range, 0–1; higher values indicate greater overlap). For each patient, the deformation field from the sequence with the highest brain Dice was retained, and the tumor mask was transformed into Haskins template space using nearest-neighbor interpolation to preserve discrete label values. All probabilistic maps and atlas-derived spatial features were computed in Haskins template space.

For quality control, registrations were visually reviewed in ascending order of brain Dice by reviewers blinded to molecular subtype and outcome. Rather than using a prespecified cutoff, the exclusion threshold was defined at a natural gap in the brain Dice distribution, below which registrations showed anatomically unacceptable alignment.

### Probabilistic MRI Mapping

Within each stratum, registered binary tumor masks were averaged voxel-wise, so that each voxel represents the proportion of patients in that stratum whose tumor involved that location. Age strata were 0–2, 2–5, and 5–18 years; molecular strata were *KIAA1549–BRAF* fusion, *BRAF* V600E, and non-*BRAF*; EOR strata were GTR, STR, and biopsy/no-surgery; and sex strata were female and male. Patients without available molecular subtype were excluded from molecular subtype maps. Patients with a *KIAA1549–BRAF* fusion were assigned to the fusion group regardless of co-occurring alterations, whereas patients with *BRAF* V600E without a *KIAA1549–BRAF* fusion were assigned to the *BRAF* V600E group. The remaining patients with available molecular data comprised the non-*BRAF* group. Within the non-*BRAF* group, eight molecular categories were evaluated: molecularly tested with no listed alteration, other MAPK alteration, neurofibromatosis type 1 (NF1), receptor tyrosine kinase (RTK), FGFR, CDKN2A/B, *IDH*, and *MYB/MYBL1*. Patients with more than one listed non-*BRAF* alteration contributed to each applicable category using multi-hot assignment.

For each stratum, a voxel-wise Welch t statistic compared tumor occurrence with that of the remaining patients in the corresponding analysis set, showing the direction and magnitude of spatial differences.

### Regional Tumor Localization and Atlas-Derived Spatial Features

Patient-level tumor localization was quantified in the Haskins template space by measuring tumor overlap with the 107-region Haskins parcellation. Atlas-derived spatial features included regional tumor overlap and minimum tumor-to-region distance for each of the 107 Haskins regions (n = 214); tumor overlap and minimum distance for each of the 14 prespecified anatomic macroregions (n = 28); summary measures of tumor involvement across 30 regions designated as eloquent (n = 7), those in which tumor involvement carries a high risk of neurological deficit; and laterality and midline features (n = 9). Eloquent regions were designated based on the grade III eloquence definition of Sawaya et al. [20], applied at the resolution of the parcellation. Regional tumor overlap was defined as the proportion of each region occupied by tumor, and tumor voxels not assigned to a Haskins label were excluded from regional overlap calculations. Minimum tumor-to-region distance was defined as the Euclidean distance from the tumor to each region, set to zero where the two intersect. In addition, 14 tumor morphometric features captured tumor shape and the volumes of enhancing, nonenhancing, and cystic components. Together, these comprised 272 atlas-derived spatial and tumor morphometric features for each patient. Detailed definitions are provided in the Supplementary Methods, the mapping of Haskins regions to macroregions in Supplementary Table S1, the complete feature set in Supplementary Table S2, and the eloquent regions in Supplementary Table S3. To quantify the spatial patterns observed in the probabilistic maps, regional tumor overlap was compared across age, molecular subtype, EOR, and sex using the Kruskal–Wallis test, followed by group-versus-rest Mann–Whitney U tests with rank-biserial correlation as the effect size [21]. Benjamini–Hochberg correction was applied within each variable and spatial resolution [22], with *q* < 0.05 considered statistically significant.

### Classification of EOR

The Discovery (training/validation) partition, on which the classifiers were tuned and fit, and the Replication (testing, unseen during model development) partition, scored once, were those of the previously published clinico-imaging risk stratification models [6,7]. A two-level classification approach was implemented by training two calibrated gradient-boosted classifiers (XGBoost) [23] using nested cross-validation (5 inner, 5 outer folds) on the Discovery partition: one distinguishing GTR from STR and biopsy/no-surgery, and one distinguishing biopsy/no-surgery from STR. Candidate predictors were the atlas-derived spatial features, tumor morphometric features, and clinical covariates age, sex, and NF1 status. Within each outer training fold, continuous features were clustered by Spearman rank correlation at |ρ| = 0.70 with complete linkage, and the feature with the largest rank-biserial correlation was retained from each cluster; the selection procedure is given in the Supplementary Methods. Imputation and standardization were fit inside every training fold. The classifiers were then retuned on the complete Discovery partition and scored once in the Replication partition, with probabilities calibrated by Platt scaling on out-of-fold predictions. Shapley additive explanations (SHAP) analysis was used to assess feature contributions [24], and the hyperparameter search spaces are provided in the Supplementary Methods. Model performance was summarized by the area under the receiver operating characteristic curve (AUC) with a 2,000-sample percentile bootstrap 95% confidence interval (CI), the Brier score, and the calibration slope.

### Clinico-Imaging Risk Stratification

To test whether the 107 atlas-derived regional overlap features could be used in place of the manually assigned tumor location variable, a categorical variable with nine values (basal ganglia, brainstem, cerebellar, intraventricular, lobar, multi-regional, suprasellar, thalamus, or other) [6], we substituted them for that variable within two previously described machine/deep learning predictive clinico-imaging models for risk stratification of pLGGs: the clinicoradiomic model, which combines clinical predictors with multiparametric radiomic features [6], and the clinico-ResNet model, which combines the same clinical predictors with deep learning image features [7]. Each of the four resulting feature sets was fit with a penalized Cox model [25]. Because the substitution introduces 107 predictors in place of one, the regional overlap feature block carried its own penalty factor, selected within the Discovery cross-validation. Every other predictor, the outcome definition, the outer cross-validation folds, and the partition assignment recorded in the clinical table were held fixed. Model performance was evaluated using the Harrell [26] and Uno [27] concordance indices (C-indices), giving four paired comparisons: two imaging feature sets × two indices. Cross-validation used 10 outer and 10 inner folds, identical across all four feature sets, so the per-fold values are paired. Discovery C-indices are reported as the mean over the 10 outer folds with a 95% CI across those folds, and Replication C-indices with a 500-sample percentile bootstrap 95% CI. Paired outer-fold differences were assessed using the corrected resampled paired t test [28].

No formal sample size calculation was performed, and all patients meeting the inclusion criteria were analyzed. Statistical analyses were performed in Python 3.12.12 (Python Software Foundation) with the packages listed in the Supplementary Methods, and *P* < 0.05 was considered statistically significant.

## Results

### Cohort Characteristics and Atlas Registration Performance

The final cohort comprised 349 patients (median age, 9.2 years [interquartile range (IQR), 4.9–12.7 years]; male, n = 193 [55.3%]) (Table 1). Molecular subtype was available in n = 287 (82.2%): *KIAA1549–BRAF* fusion (n = 139 [48.4%]), *BRAF* V600E (n = 43 [15.0%]), and non-*BRAF* (n = 105 [36.6%]); subtype was unavailable in n = 62 (17.8%). EOR was GTR (n = 177 [50.7%]), STR (n = 98 [28.1%]), or biopsy/no-surgery (n = 74 [21.2%]). Median follow-up, estimated using the reverse Kaplan–Meier method, was 44.9 months (IQR, 22.4–61.4 months), and n = 112 (32.1%) experienced a PFS event. PFS by EOR is shown in Supplementary Analysis S1 and Supplementary Figure S1.

**Table 1.** Characteristics of the 349 Patients with Pediatric Low-Grade Glioma.

| Characteristic | Value |
| --- | --- |
| Total patients, n | 349 |
| Age at imaging, y, median (IQR) | 9.2 (4.9, 12.7) |
| 0–2 y, n (%) | 21 (6.0) |
| 2–5 y, n (%) | 71 (20.3) |
| 5–18 y, n (%) | 257 (73.6) |
| Sex, male, n (%) | 193 (55.3) |
| NF1 status, n (%) | 19 (5.4) |
| Molecular subtype known, n (%) | 287 (82.2) |
| *KIAA1549–BRAF* fusion, n (%)* | 139 (48.4) |
| *BRAF* V600E, n (%)* | 43 (15.0) |
| Non-*BRAF*, n (%)* | 105 (36.6) |
| Extent of resection |  |
| GTR, n (%) | 177 (50.7) |
| STR, n (%) | 98 (28.1) |
| Biopsy/no-surgery, n (%) | 74 (21.2) |
| Driving MRI sequence, T1 / FLAIR, n | 286 / 63 |
| Median brain Dice (IQR) | 0.986 (0.984, 0.987) |
| PFS events, n (%) | 112 (32.1) |
| Median follow-up, mo (IQR) | 44.9 (22.4, 61.4) |

*Note: Data are for the single analytic cohort of 349 patients retained after registration quality control (11 of 360 eligible patients excluded for registration quality-control failure). IQR = interquartile range, NF1 = neurofibromatosis type 1, PFS = progression-free survival, FLAIR = fluid-attenuated inversion recovery, GTR = gross total resection/near total resection, STR = subtotal resection, MRI = magnetic resonance imaging, T1 = T1-weighted. \*Subtype percentages are of the 287 patients with a known molecular subtype.*

The selected registrations for all 360 patients underwent visual quality control, and 11 were excluded because of anatomically unacceptable alignment, leaving 349 patients for analysis. Among retained registrations, median brain Dice was 0.986 (IQR, 0.984–0.987). The brain Dice threshold of 0.92 corresponded to a natural gap in the distribution. The lowest retained registration had a brain Dice of 0.927, whereas the highest excluded registration had a brain Dice of 0.901. Qualitative review confirmed anatomically coherent alignment across age groups, including children aged 0–2 years (n = 21), the group most susceptible to misregistration when an adult template is used.

### Probabilistic MRI Mapping

#### Age

Tumor distribution varied significantly with age (Figure 2; *P* < 0.001). Probabilistic maps showed greater involvement of the optic pathway and suprasellar region and deep midline structures in children aged 0–2 years (n = 21), whereas tumors in children aged 5–18 years (n = 257) showed less involvement of these regions and greater cerebellar involvement. Regional analysis identified age-related differences in 9 of 14 anatomic macroregions, with the strongest difference in the optic pathway and suprasellar region (Kruskal–Wallis *H* = 32.7; *q* = 1.1 × 10⁻⁶). Compared with the remainder of the cohort, children aged 0–2 years had greater optic pathway and suprasellar involvement (rank-biserial *r* = 0.48; *q* = 4.3 × 10⁻⁶), whereas children aged 5–18 years had less involvement (*r* = −0.20; *q* = 7.2 × 10⁻⁴). Children aged 2–5 years (n = 71) showed an intermediate spatial distribution, with no individual macroregion differing significantly from the remainder of the cohort after multiple-comparison correction.

**Figure 2.**
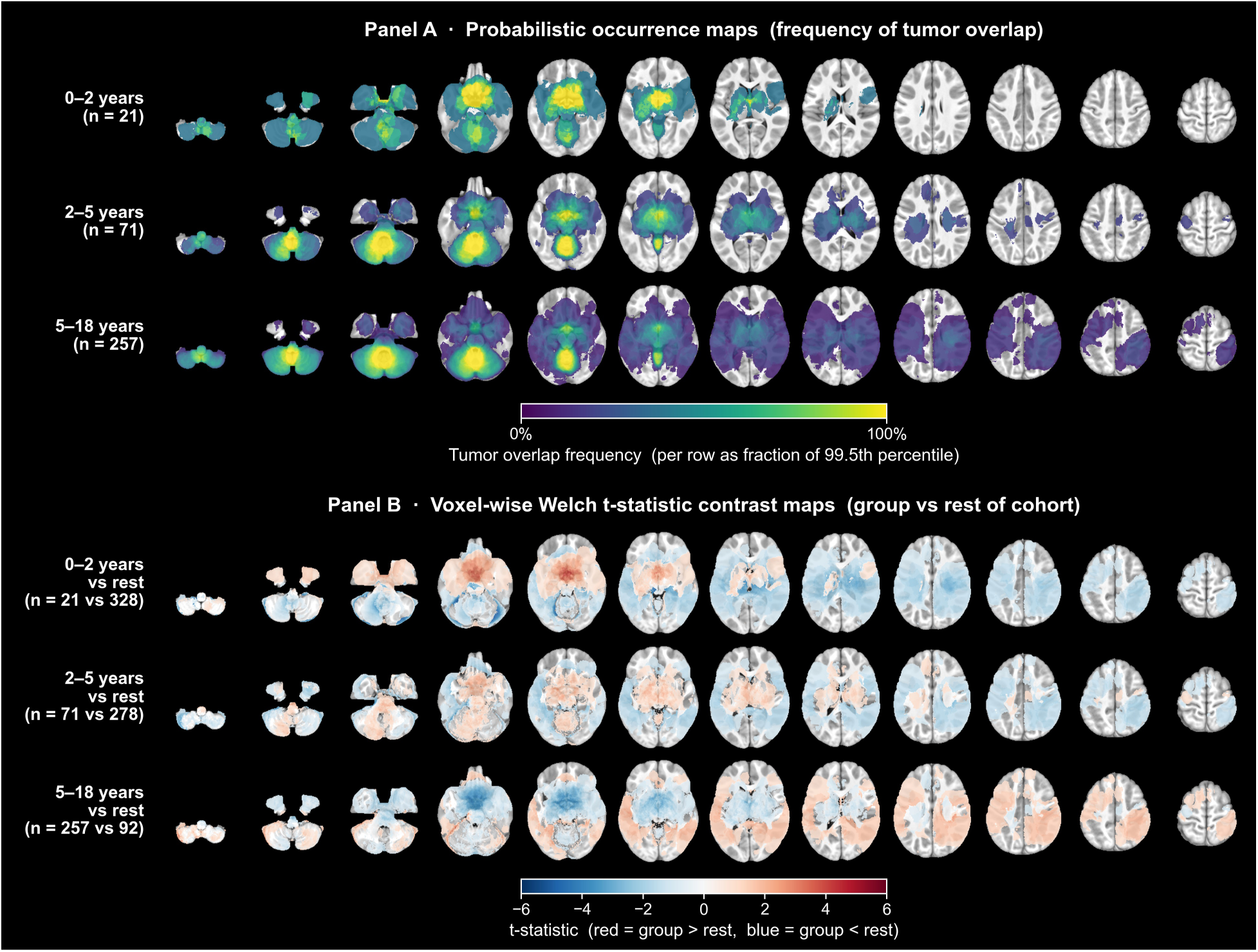
Age-stratified spatial maps in Haskins template space. (A) Probabilistic maps for each of the three strata (0–2 years, n = 21; 2–5 years, n = 71; 5–18 years, n = 257), shown as 12 axial slices inferior to superior on the viridis scale, which is scaled within each row to that row’s 99.5th percentile of overlap frequency, so that color is comparable within a row but not between rows. (B) Voxel-wise Welch t statistic maps comparing each age stratum against the rest of the cohort on a red–blue diverging scale, red indicating greater tumor probability in the named stratum. Tumor distribution shifts from optic pathway, suprasellar, and deep midline enrichment in the youngest stratum to cerebellar predominance in older children and adolescents. Region-level testing over 14 macroregions found 9 differing across strata, led by the optic pathway and suprasellar region (Kruskal–Wallis *H* = 32.7, *q* = 1.1 × 10⁻⁶), with greater involvement in the 0–2-year stratum (rank-biserial *r* = 0.48, *q* = 4.3 × 10⁻⁶) and less involvement in the 5–18-year stratum (*r* = −0.20, *q* = 7.2 × 10⁻⁴). These age–location associations were not adjusted for molecular subtype.

#### Molecular Subtype

Molecular subtypes showed distinct tumor distributions (Figure 3A, 3B). *KIAA1549–BRAF* fusion tumors (n = 139) showed greater cerebellar involvement (rank-biserial *r* = 0.53; *q* < 1 × 10⁻¹¹). *BRAF* V600E tumors (n = 43) showed less cerebellar involvement (*r* = −0.44; *q* = 2 × 10⁻⁵) and greater temporal cortical involvement (*r* = 0.30; *q* = 0.003). Non-*BRAF* tumors (n = 105) showed less cerebellar involvement (*r* = −0.32; *q* = 2 × 10⁻⁵) and greater frontal cortical (*r* = 0.23; *q* = 0.001) and deep gray involvement (*r* = 0.19; *q* = 0.0083). Across the three molecular groups, 11 of 14 macroregions differed, with the largest differences in the cerebellum (Kruskal–Wallis *H* = 65.0; *q* < 1 × 10⁻¹¹) and brainstem (*H* = 45.9; *q* < 1 × 10⁻⁶). Within the non-*BRAF* group, additional maps showed prominent optic pathway and hypothalamic involvement in NF1-associated tumors, frontal involvement in IDH-mutant tumors, and temporal and limbic involvement in MYB/MYBL1 tumors (Figure 3C, 3D).

**Figure 3.**
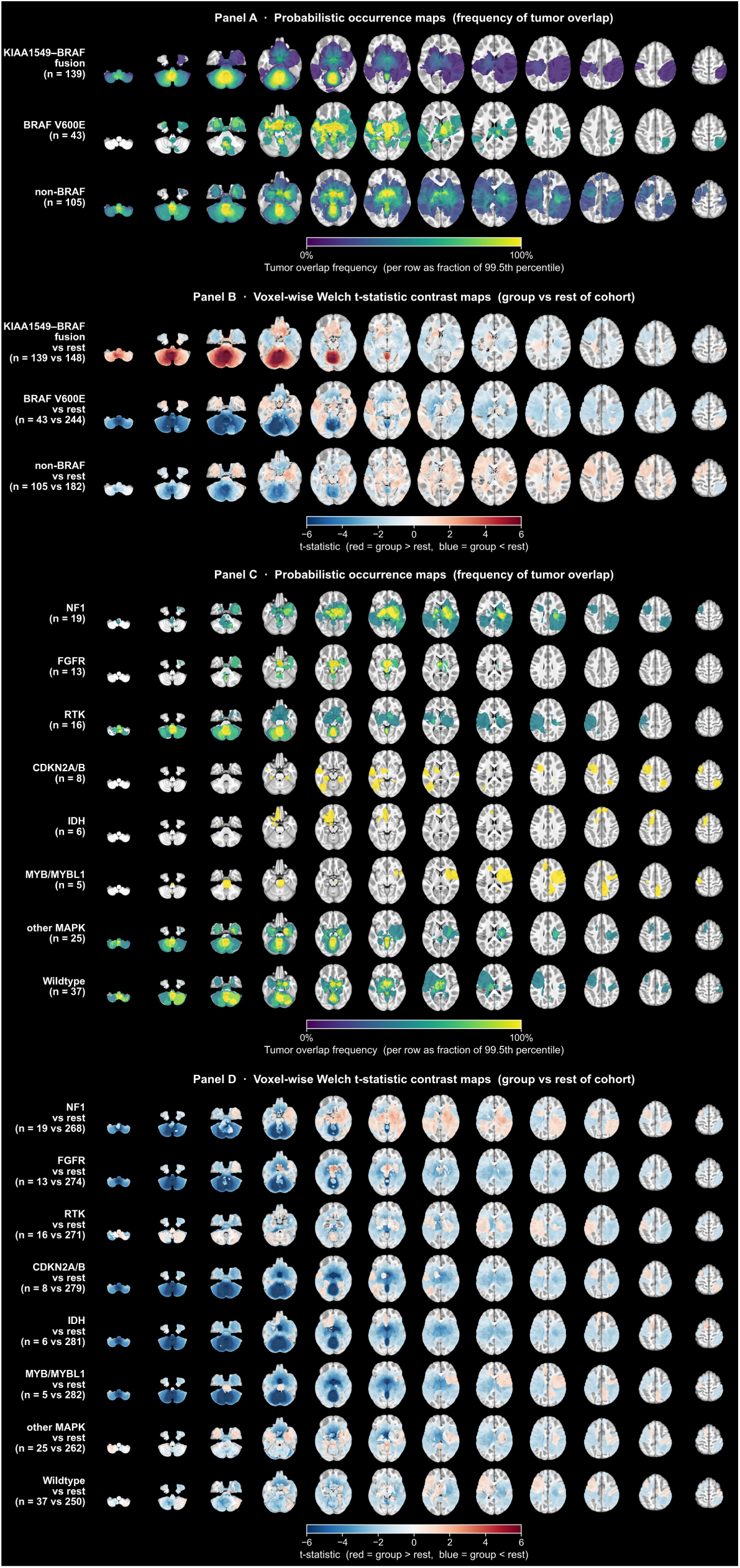
Molecular-subtype spatial maps. (A, B) Three-category maps (n = 287 with known molecular subtype). (A) Probabilistic maps for *KIAA1549–BRAF* fusion (n = 139), *BRAF* V600E (n = 43), and non-*BRAF* (n = 105), viridis scale, scaled within each row to that row’s 99.5th percentile of overlap frequency. (B) Voxel-wise Welch t statistic maps comparing each subtype against the rest of the annotated cohort, red–blue diverging scale. Eleven of 14 macroregions differed across subtypes, led by the cerebellum (Kruskal–Wallis *H* = 65.0, *q* < 1 × 10⁻¹¹). Cerebellar involvement is greater in *KIAA1549–BRAF* fusion tumors (rank-biserial *r* = 0.53, *q* < 1 × 10⁻¹¹) and less in *BRAF* V600E tumors (*r* = −0.44, *q* = 2 × 10⁻⁵), which show greater temporal cortical involvement (*r* = 0.30, *q* = 0.003). The three categories in (A, B) are assigned hierarchically. Patients with a listed *KIAA1549–BRAF* fusion are assigned to *KIAA1549–BRAF* regardless of co-occurring alterations, patients with *BRAF* V600E and no *KIAA1549–BRAF* fusion enter the *BRAF* V600E group, and the remainder with any listed non-*BRAF* alteration or wildtype status form the non-*BRAF* group. (C, D) Non-*BRAF* subtype breakdown, shown as in (A, B), using multi-hot classification: NF1 (n = 19, germline and somatic combined), *FGFR* (n = 13), RTK (n = 16), *CDKN2A/B* (n = 8), *IDH* (n = 6), *MYB/MYBL1* (n = 5), other MAPK (n = 25), and wildtype (n = 37). “Wildtype” denotes patients molecularly tested with no listed alteration. Patients with more than one listed alteration contribute to each of their alteration maps, and given the small subgroup sizes these maps are presented as exploratory anatomic descriptions rather than confirmatory comparisons. NF1 tumors show prominent optic pathway and hypothalamic involvement, *IDH* tumors frontal involvement, and *MYB/MYBL1* tumors temporal and limbic involvement. Overlays are shown on axial slices of the Haskins pediatric template, and 62 patients lacked molecular annotation and are excluded. *CDKN2A/B* = cyclin-dependent kinase inhibitor 2A/B, *FGFR* = fibroblast growth factor receptor, *IDH* = isocitrate dehydrogenase, MAPK = mitogen-activated protein kinase, NF1 = neurofibromatosis type 1, RTK = receptor tyrosine kinase.

#### Extent of Resection

Tumor distribution differed across EOR categories (Figure 4). Tumors with GTR (n = 177) were concentrated in the cerebellar hemispheres and supratentorial lobar cortex, whereas tumors with biopsy/no-surgery (n = 74) were concentrated in the thalamus, deep midline, and suprasellar region; tumors with STR (n = 98) showed an intermediate distribution. Thirteen of 14 macroregions differed across EOR categories, with the largest difference in the deep gray nuclei (Kruskal–Wallis *H* = 72.4; *q* = 2.7 × 10⁻¹⁵). Deep gray involvement was lower in the GTR (rank-biserial *r* = −0.48; *q* = 8.6 × 10⁻¹⁶) and greater in the biopsy/no-surgery (*r* = +0.28; *q* = 1.3 × 10⁻⁴) and STR (*r* = +0.36; *q* = 6.3 × 10⁻⁸) groups. Cerebellar involvement was greater in the GTR (*r* = +0.37; *q* = 6.7 × 10⁻⁹) and lower in the biopsy/no-surgery (*r* = −0.33; *q* = 1.6 × 10⁻⁵) group. Optic pathway and suprasellar involvement was lower in the GTR (*r* = −0.31; *q* = 2.7 × 10⁻¹¹) and greater in the STR (*r* = +0.25; *q* = 1.2 × 10⁻⁶) and biopsy/no-surgery (*r* = +0.16; *q* = 0.0045) groups. Ventricular involvement was greater in the STR (*r* = +0.25; *q* = 5.4 × 10⁻⁴) and lower in the biopsy/no-surgery (*r* = −0.27; *q* = 5.9 × 10⁻⁴) group. Selected spatial, morphometric, and clinical features associated with EOR are shown in Table 2. At the 107-region resolution, 49 regions differed across EOR categories. Sex-stratified maps are shown in Supplementary Analysis S2 and Supplementary Figure S2.

**Figure 4.**
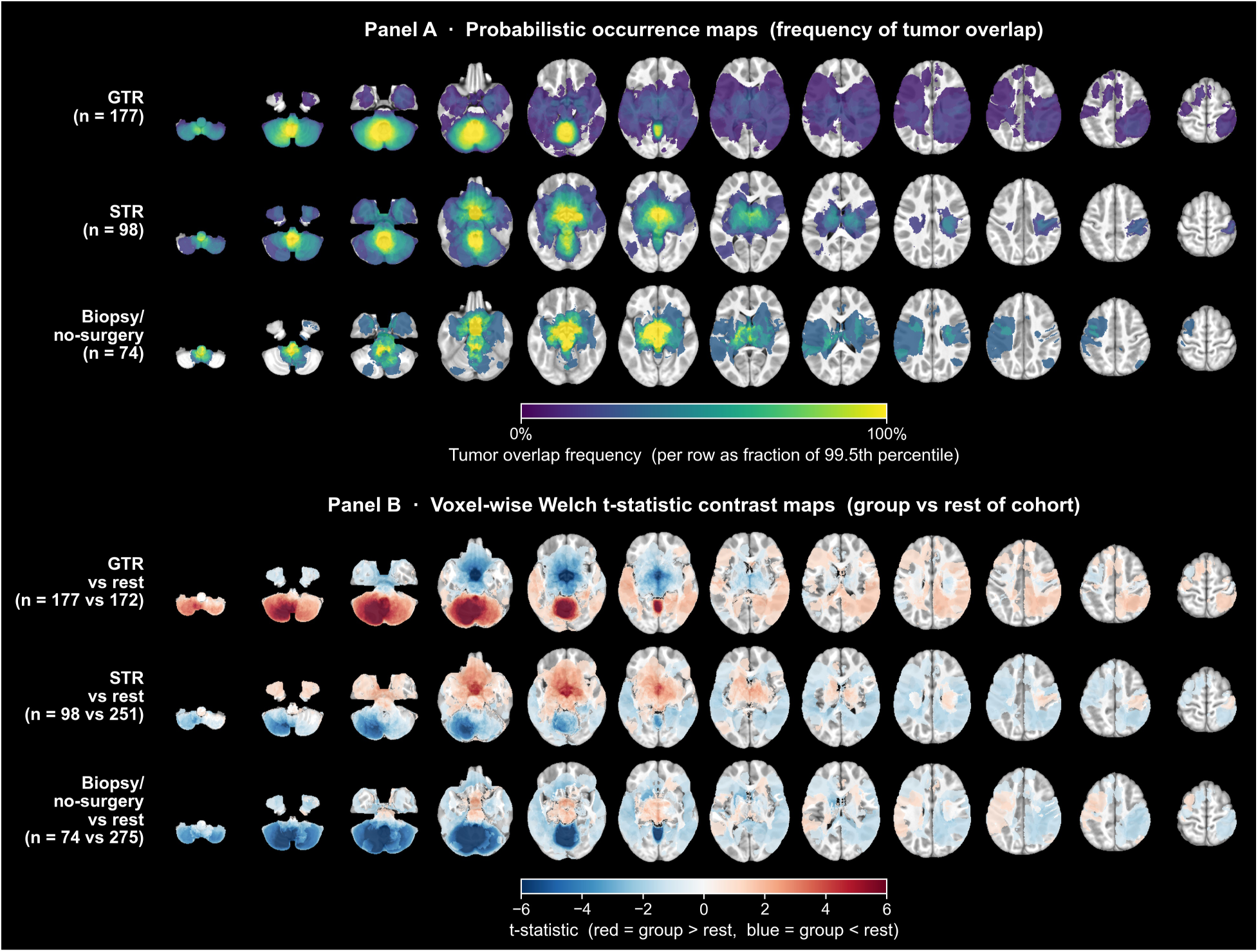
Extent-of-resection spatial distribution. (A) Probabilistic maps by category: GTR (n = 177), STR (n = 98), and biopsy/no-surgery (n = 74), viridis scale, scaled within each row to that row’s 99.5th percentile of overlap frequency. (B) Voxel-wise Welch t statistic maps comparing each resection category against the rest of the cohort, red–blue diverging scale. Overlays are shown on axial slices of the Haskins pediatric template. Biopsy/no-surgery probability concentrates in the deep midline, thalamus, and suprasellar region, and its lower probability in the cerebellar hemispheres produces the largest voxel-wise t statistic among the resection categories (peak |t| = 12.1), whereas GTR probability predominates in the cerebellar hemispheres and lobar cortex. Thirteen of 14 macroregions differed across categories, led by the deep gray nuclei (Kruskal–Wallis *H* = 72.4, *q* = 2.7 × 10⁻¹⁵), with lower involvement in GTR (rank-biserial *r* = −0.48, *q* = 8.6 × 10⁻¹⁶) and greater involvement in biopsy/no-surgery (*r* = +0.28, *q* = 1.3 × 10⁻⁴). The values reported here were computed in the full cohort (n = 349) with correction across the 14 anatomic macroregions, whereas those in Table 2 come from the decorrelated feature-selection analysis in the Discovery partition (n = 273), so the values in the two are not directly comparable. GTR = gross total resection/near total resection, STR = subtotal resection.

**Table 2.** Selected Spatial, Morphometric, and Clinical Features Associated with Extent of Resection.

| Region or feature | Contrast | Effect size | *q* value |
| --- | --- | --- | --- |
| Deep gray nuclei | GTR vs rest | $r = -0.44$ (95% CI: $-0.55, -0.33$) | $7.6 \times 10^{-11}$ |
| Cerebellum | GTR vs rest | $r = +0.32$ (95% CI: $+0.20, +0.45$) | $6.0 \times 10^{-6}$ |
| Deep gray nuclei | Biopsy/no-surgery vs rest | $r = +0.23$ (95% CI: $+0.08, +0.38$) | 0.013 |
| Deep gray nuclei | STR vs rest | $r = +0.36$ (95% CI: $+0.23, +0.49$) | $9.4 \times 10^{-6}$ |
| Cerebellum | Biopsy/no-surgery vs rest | $r = -0.31$ (95% CI: $-0.44, -0.19$) | 0.0014 |
| Tumor surface-to-volume ratio | Biopsy/no-surgery vs rest | $r = +0.47$ (95% CI: $+0.34, +0.60$) | $1.4 \times 10^{-5}$ |
| Neurofibromatosis type 1 status | Biopsy/no-surgery vs rest | OR = 6.02 (95% CI: 2.24, 16.16) | 0.0041 |

*Note: Values are representative decorrelated spatial and morphometric features for each extent-of-resection comparison. $r$ = rank-biserial correlation, OR = odds ratio, $q$ = Benjamini–Hochberg false-discovery-rate–corrected value, CI = confidence interval, GTR = gross total resection/near total resection, STR = subtotal resection. Effect direction is relative to each comparison's positive class, biopsy/no-surgery for biopsy/no-surgery vs rest, STR for STR vs rest, and GTR for GTR vs rest. Feature definitions are provided in the Supplementary Methods. These values come from the decorrelated feature-selection analysis described in the Methods, computed in the Discovery partition ( $n = 273$ ) with correction across the retained features within each comparison, and are not directly comparable with the macroregion values reported in the Results and in the Figure 4 legend, which were computed in the full cohort ( $n = 349$ ) with correction across the 14 anatomic macroregions.*

### Classification of EOR

The regional analyses identified associations between tumor location and EOR. As a secondary patient-level analysis, we evaluated whether atlas-derived spatial, tumor morphometric, and clinical features could distinguish GTR from STR and biopsy/no-surgery. A calibrated XGBoost classifier was fit on the Discovery partition (n = 273) with 60 selected features and scored once on the Replication partition (n = 76). The AUC was 0.852 (95% CI: 0.807, 0.895) on pooled Discovery out-of-fold predictions and 0.864 (95% CI: 0.772, 0.944) in the Replication partition (sensitivity, 92% [35 of 38]; specificity, 66% [25 of 38]). Brier scores were 0.157 and 0.144, and calibration slopes were 1.05 and 1.12 in the Discovery and Replication partitions, respectively. Summed by macroregion, mean absolute SHAP values ranked the deep gray nuclei first (0.82), followed by the parietal (0.43) and temporal (0.39) cortices and the ventricular structures and cerebrospinal fluid (0.26), consistent with the regional analysis and the probabilistic maps. The largest individual contributions were distance to the left ventral diencephalon, cystic tumor volume, and midline fraction (Figure 5). A second classifier distinguishing biopsy/no-surgery from STR achieved an AUC of 0.652 (95% CI: 0.556, 0.741) in the Discovery partition and 0.855 (95% CI: 0.717, 0.958) in the Replication partition (Supplementary Analysis S3, Supplementary Figure S3).

**Figure 5.**
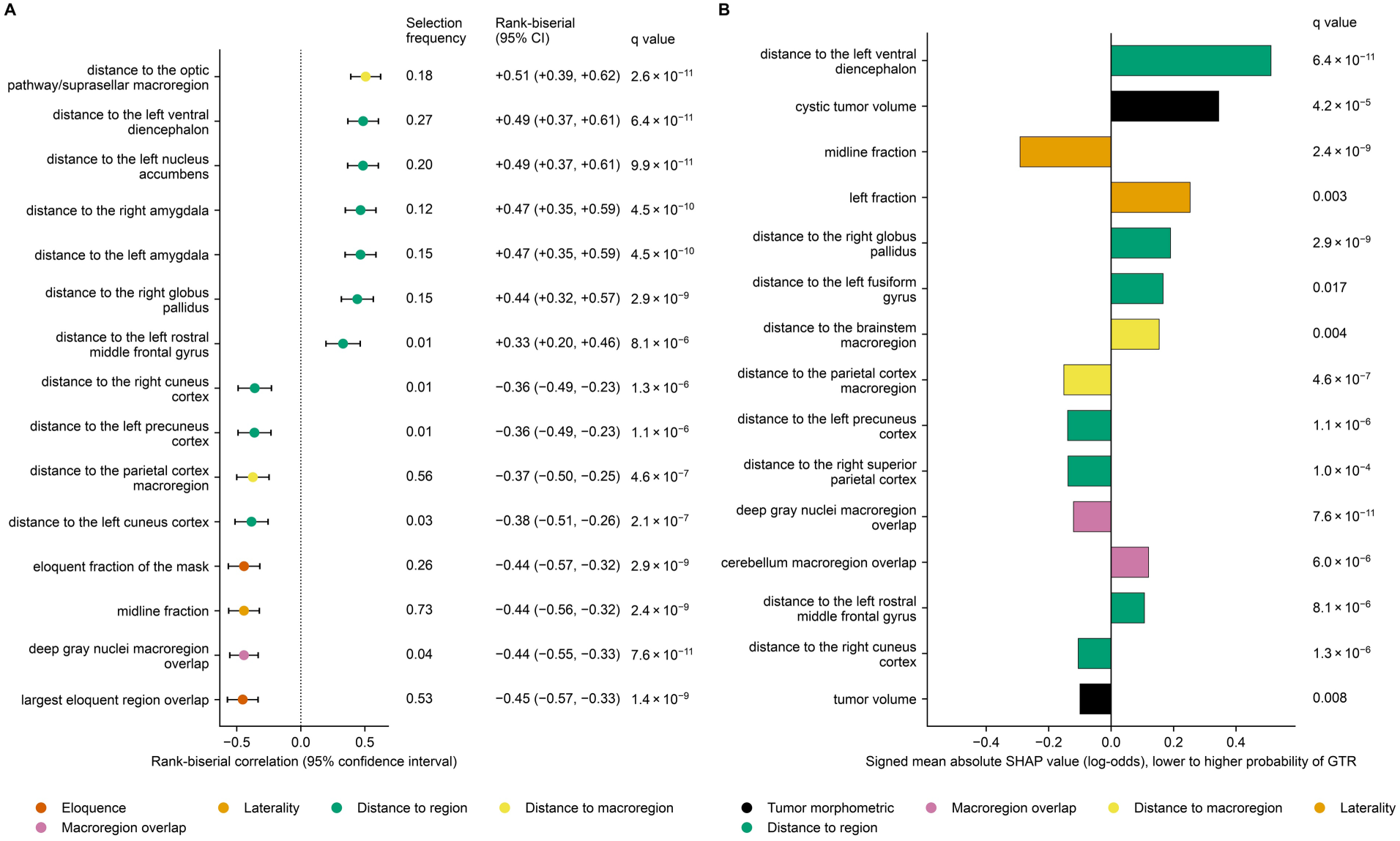
Atlas-derived spatial features associated with EOR. (A) The 15 largest univariate effects distinguishing GTR from STR or biopsy/no-surgery in the Discovery partition, shown as rank-biserial correlations with 95% CIs. Features are grouped by feature family and annotated with the corresponding selection frequency, effect size, 95% CI, and false-discovery-rate–corrected *q* value; the selection frequency is a descriptive stability statistic defined in the Supplementary Methods and does not itself select features. Features represent the decorrelated cluster representatives described in the Methods. (B) Signed mean absolute SHAP values for the classifier distinguishing GTR from STR or biopsy/no-surgery. Bars to the right indicate features associated with a higher predicted probability of GTR, and bars to the left indicate features associated with a lower predicted probability of GTR, and each feature is annotated with its corresponding univariate *q* value. CI = confidence interval, EOR = extent of resection, GTR = gross total resection/near total resection, SHAP = Shapley additive explanations, STR = subtotal resection.

### Clinico-Imaging Risk Stratification

The manually assigned tumor location variable in the two clinico-imaging models was replaced with the 107 atlas-derived regional overlap features, while the imaging features, clinical predictors, and outer cross-validation folds remained unchanged (Figure 6). The Discovery partition included 273 patients with 81 PFS events and the Replication partition included 76 patients with 31 events. In the clinicoradiomic penalized Cox model, the Discovery Harrell C-index was 0.735 (95% CI: 0.668, 0.801) with the manually assigned tumor location variable and 0.740 (95% CI: 0.684, 0.796) with the 107 atlas-derived regional overlap features, a paired difference of +0.006 (95% CI: −0.042, +0.053; corrected resampled paired t test, *P* = 0.80), and the Discovery Uno C-index was 0.722 (95% CI: 0.654, 0.790) and 0.732 (95% CI: 0.676, 0.787), a paired difference of +0.010 (95% CI: −0.038, +0.057; *P* = 0.65); in the Replication partition, the corresponding Harrell C-index values were 0.693 (95% CI: 0.588, 0.793) and 0.697 (95% CI: 0.596, 0.792) and the Uno C-index values were 0.696 (95% CI: 0.593, 0.800) and 0.694 (95% CI: 0.591, 0.791). In the clinico-ResNet penalized Cox model, the Discovery Harrell C-index was 0.707 (95% CI: 0.637, 0.777) and 0.729 (95% CI: 0.666, 0.792), a paired difference of +0.022 (95% CI: −0.021, +0.066; *P* = 0.27), and the Discovery Uno C-index was 0.689 (95% CI: 0.614, 0.763) and 0.720 (95% CI: 0.660, 0.780), a paired difference of +0.031 (95% CI: −0.010, +0.072; *P* = 0.12); in the Replication partition, the corresponding Harrell C-index values were 0.693 (95% CI: 0.589, 0.792) and 0.691 (95% CI: 0.584, 0.795) and the Uno C-index values were 0.703 (95% CI: 0.600, 0.798) and 0.693 (95% CI: 0.583, 0.792). All four Discovery differences were positive, every confidence interval included zero, and none of the four paired differences was statistically significant (smallest *P* = 0.12), indicating that replacing the manually assigned tumor location variable with the atlas-derived regional overlap features did not significantly alter discrimination in the evaluated models.

**Figure 6.**
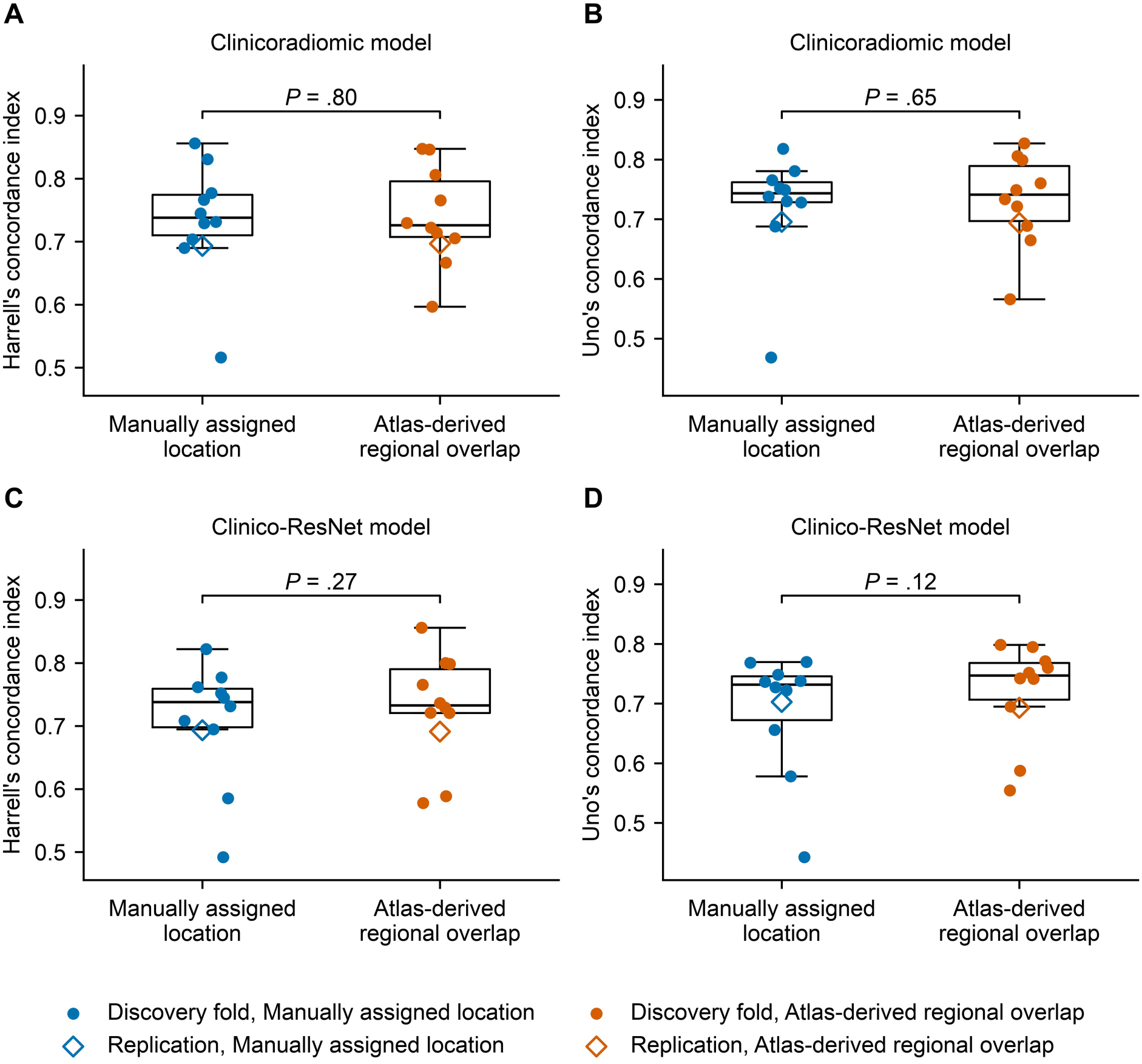
Atlas-derived regional overlap features in clinico-imaging risk stratification models, replacing a manually assigned category with features computed automatically from the tumor mask. The manually assigned tumor location variable was replaced with the 107 atlas-derived regional overlap features, while the clinical predictors, imaging features, and outer cross-validation folds remained unchanged. (A, B) Clinicoradiomic models, Harrell and Uno C-indices. (C, D) Clinico-ResNet models, the same two indices. Each panel shows paired outer-fold model performance for the manually assigned tumor location variable and the 107 atlas-derived regional overlap features on identical folds. Boxes show the distribution across outer folds, individual points show the corresponding fold values, and the open diamond shows model performance in the Replication partition. Corrected resampled paired t test *P* values are shown for each comparison. None of the four paired differences was statistically significant (smallest *P* = 0.12). C-index = concordance index, ResNet = residual neural network.

## Discussion

In this study of 349 children with pLGG, we developed and evaluated a reproducible pediatric atlas-based framework for spatial phenotyping, in which tumor location is represented quantitatively across the full spatial extent of the tumor rather than as a single manually assigned anatomic category. The framework combines voxel-wise probabilistic maps of tumor distribution with patient-level atlas-derived spatial features. The probabilistic maps and regional analyses showed differences in tumor distribution by age, molecular subtype, and EOR that were consistent with previous studies, providing a quantitative representation of established spatial patterns. Atlas-derived spatial, tumor morphometric, and clinical features also supported classification of EOR. Replacing the manually assigned tumor location variable with the 107 atlas-derived regional overlap features, computed automatically from the tumor mask, did not significantly alter discrimination in the evaluated clinico-imaging risk stratification models.

Prior probabilistic mapping in adult glioma and meningioma has demonstrated the value of quantitative spatial analysis [8–10]. In pLGG, prior spatial probability distributions were derived in an adult reference space for molecular subtype classification [11]. In contrast, the present study provides a pediatric atlas-based framework that integrates voxel-wise probabilistic mapping with patient-level atlas-derived spatial features within the same pediatric reference space. This enables the same quantitative representation of tumor location to be examined across age, molecular subtype, and EOR and to be carried forward into patient-level analyses, including EOR classification and clinico-imaging risk stratification. The same atlas-derived spatial features were incorporated alongside different imaging and clinical feature sets, including radiomic and deep learning image features. The publicly available implementation provides a reproducible workflow for pediatric atlas-based tumor mapping and regional spatial feature extraction, facilitating application and evaluation in other pediatric neuro-oncology cohorts.

The probabilistic maps and regional analyses showed distinct spatial patterns across age, molecular subtype, and EOR. Tumors in the youngest children showed greater involvement of the optic pathway and suprasellar region and deep midline structures, whereas older children showed greater cerebellar involvement. *KIAA1549–BRAF* fusion tumors showed greater cerebellar involvement, *BRAF* V600E tumors showed greater temporal cortical involvement, and non-*BRAF* tumors showed greater frontal cortical and deep gray involvement. Across EOR categories, tumors with GTR were concentrated in the cerebellar hemispheres and supratentorial lobar cortex, whereas tumors with biopsy/no-surgery were concentrated in the thalamus, deep midline, and suprasellar region, with STR showing an intermediate distribution. These patterns were consistent with previous studies of pLGG location across age, molecular subtype, and EOR [2–5,11,29]. Their representation using both probabilistic maps and regional overlap features supports the validity of the framework for quantitative spatial phenotyping.

The patient-level analyses further demonstrated how this spatial representation can be applied beyond group-level mapping. An XGBoost model incorporating atlas-derived spatial, tumor morphometric, and clinical features distinguished GTR from STR or biopsy/no-surgery in the Replication partition, with an AUC of 0.864 (95% CI: 0.772, 0.944). Summed mean absolute SHAP values ranked the deep gray nuclei first, followed by the parietal and temporal cortices, consistent with the regional analyses and probabilistic maps. The classifier therefore extends the relationship between tumor location and EOR identified in the regional analyses to patient-level classification rather than providing independent evidence of a separate association. The analysis was intended to characterize these associations and not to determine the EOR that is safe or achievable for an individual patient. The secondary classifier distinguishing biopsy/no-surgery from STR showed less consistent and less precise performance, with AUCs of 0.652 (95% CI: 0.556, 0.741) in the Discovery partition and 0.855 (95% CI: 0.717, 0.958) in the Replication partition, indicating substantial uncertainty in distinguishing these two EOR categories.

A second patient-level application evaluated whether the 107 atlas-derived regional overlap features could be used in place of the manually assigned tumor location variable within previously published clinico-imaging risk stratification models [6,7]. None of the four paired Discovery comparisons was statistically significant, and Replication C-index values differed by 0.010 or less. These comparisons were not designed to establish equivalence or to develop a new prognostic model. The imaging features, clinical predictors, and outer cross-validation folds were held unchanged between paired comparisons. Within these paired comparisons, replacing a single manually assigned tumor location variable with regional overlap features computed automatically from the tumor mask, which represent tumor location across the full spatial extent of the tumor, did not significantly alter discrimination.

This study has limitations. The Haskins pediatric template was constructed from children aged 7–14 years [13], whereas the cohort spanned 0–18 years. Although registration quality was high and qualitative review confirmed anatomically coherent alignment across age groups, including the youngest children, pediatric templates spanning the full developmental age range would strengthen future studies. Registration may also be less accurate in tumors producing substantial mass effect or anatomic distortion, and exclusion of anatomically unacceptable registrations may underrepresent tumors with the greatest distortion. The non-*BRAF* molecular categories were relatively small and nonexclusive. Although the cohort was obtained through CBTN, most patients were from a single institution, and EOR is also determined by center and surgeon, which may contribute additional variability. EOR was recorded at the contributing institutions without a uniform residual-volume threshold or central review of postoperative MRI, so the EOR analyses characterize the recorded EOR rather than a standardized endpoint. Both patient-level analyses used internal Discovery and Replication partitions rather than independent external cohorts, limiting generalizability. Requiring analyzable preoperative MRI and high-quality tumor segmentation may also underrepresent patients with less complete imaging data.

In conclusion, this study provides a reproducible pediatric atlas-based framework for quantitative spatial phenotyping of pLGG using voxel-wise probabilistic maps and patient-level atlas-derived spatial features. Atlas-derived spatial, tumor morphometric, and clinical features supported classification of EOR, and replacing the manually assigned tumor location variable with automated atlas-derived regional overlap features did not significantly alter discrimination in the evaluated clinico-imaging risk stratification models. With external validation, the framework could provide a common quantitative representation of tumor location across the full spatial extent of the tumor for imaging and clinical studies of pLGG and other pediatric brain tumors.

## Data availability

Raw images, clinical variables, and molecular information are not publicly available to protect patient privacy and can be requested from CBTN (cbtn.org), subject to its data access approval.

## Code availability

The reproducible pediatric atlas-based framework is publicly available at https://github.com/d3b-center/plgg-atlas-phenotyping.

