## Supplementary for "Atlas-Based MRI Spatial Phenotyping in Pediatric Low-Grade Glioma"

### Supplementary Information

#### Supplementary Methods

##### Atlas-Derived Spatial and Tumor Morphometric Features

For each patient, tumor location was quantified in Haskins template space by intersecting the registered whole tumor mask with the 107-region SynthSeg+-derived parcellation in Haskins template space. Atlas-derived spatial features included regional tumor overlap and minimum tumor-to-region distance for each of the 107 Haskins regions (n = 214), tumor overlap and minimum distance for each of 14 prespecified anatomic macroregions (n = 28), summary measures of tumor involvement across 30 regions designated as eloquent (n = 7), and laterality and midline features (n = 9). Regional tumor overlap was defined as the proportion of each region occupied by tumor. Tumor voxels not assigned to a Haskins region were excluded from regional overlap calculations.

For each patient, minimum tumor-to-region distance was calculated in Haskins template space using a Euclidean distance transform. Distance was defined as the minimum Euclidean distance between the tumor mask and each Haskins region and was set to zero when the tumor intersected that region. Minimum distance was also summarized for each of the 14 macroregions as the minimum distance across the Haskins regions comprising that macroregion.

Seven summary measures quantified tumor involvement across 30 Haskins regions designated as eloquent. Nine additional features quantified tumor laterality and midline involvement. Detailed definitions of the eloquent region, laterality, and midline features are given below.

In addition, 14 tumor morphometric features quantified tumor shape and the volumes of enhancing, nonenhancing, and cystic components. Together, the atlas-derived spatial and tumor morphometric feature set comprised 272 features per patient. Clinical variables used in the EOR classifier, including age, sex, and neurofibromatosis type 1 (NF1) status, were considered separately and were not included in the 272-feature total.

For the EOR feature analyses, continuous features were clustered using Spearman rank correlation with a cutoff of |ρ| = 0.70 and complete linkage. Within each cluster, the feature with the largest rank-biserial correlation for the corresponding EOR comparison was retained. Binary and clinical features were retained individually. Retained continuous features were compared using the Mann–Whitney U test, with rank-biserial correlation as the effect size, and binary features using the Fisher exact test, with the odds ratio as the effect size. This procedure reduced redundancy among highly correlated features before subsequent analyses. Benjamini–Hochberg correction was applied across the retained features within each EOR comparison. Selection frequency, reported alongside each retained feature in Figure 5A, is a separate descriptive statistic: over 25 complementary half-sample pairs, an L1-penalized logistic regression was fit at each of three penalty strengths with a random per-feature reweighting, and the selection frequency is the fraction of those 150 fits in which the feature carried a nonzero coefficient. It describes stability and does not itself select features.

##### 107-Region Haskins Parcellation and 14 Anatomic Macroregions

The Haskins pediatric parcellation contains 107 cortical, subcortical, ventricular, and midline regions. These comprise 68 cortical regions, with 34 in each hemisphere, together with 39 subcortical, ventricular, and midline regions.

For regional analyses and interpretation, the 107 Haskins regions were grouped a priori into 14 anatomic macroregions: brainstem; cerebellum; corpus callosum; cingulate cortex; frontal cortex; insular cortex; occipital cortex; parietal cortex; temporal cortex; deep gray nuclei; limbic and medial temporal structures; optic pathway and suprasellar region; vascular structures; and ventricular structures and cerebrospinal fluid (CSF). The complete mapping of the 107 Haskins regions to the 14 macroregions is provided in Supplementary Table S1. The parcellation does not separately label the optic nerves, optic tracts, hypothalamus, or pituitary, so the optic pathway and suprasellar region comprises the optic chiasm label and the hypothalamus is grouped with the deep gray nuclei within the ventral diencephalon label.

##### Regional Tumor Overlap and Minimum Tumor-to-Region Distance

Regional tumor overlap was calculated as the proportion of each Haskins region occupied by tumor. Tumor voxels falling outside the labeled parcellation were excluded from regional overlap calculations. Macroregion tumor overlap was calculated as the pooled fraction, the tumor voxels within a macroregion's constituent regions divided by the combined voxels of those regions; this pooled fraction differs from the mean of the constituent regional overlaps, and it is the pooled fraction that every macroregion statistic in this study uses.

Minimum tumor-to-region distance was calculated between the whole tumor mask and each of the 107 Haskins regions in template space. Distances were derived from a single exact Euclidean distance transform of the complement of the tumor mask, set to zero when the tumor intersected the corresponding region, and capped at 200 voxels. One template voxel measures 1 mm in each direction, so distances are reported in millimeters. Minimum distances for the 14 macroregions were calculated as the minimum distance across the constituent Haskins regions.

These measures provided complementary information regarding direct regional tumor involvement and proximity to anatomically defined regions not directly involved by tumor.

##### Eloquent Regions

Tumor involvement of eloquent anatomy was summarized across 30 prespecified Haskins regions. Seven patient-level features summarized the extent and distribution of tumor involvement across these regions. The Haskins regions included in the eloquent set are listed in Supplementary Table S3, and the seven summary measures are defined below.

Write T for the set of tumor voxels, Rₛ for the voxels of Haskins region s, dₛ for the minimum tumor-to-region distance defined above, and ℓₛ for the regional tumor overlap of region s. Let E be the set of 30 eloquent regions and V = T ∩ (∪ₛ₊ₑ Rₛ) the tumor voxels within them. The seven summary features are the number of regions overlapped, |{s ∈ E : T ∩ Rₛ ≠ ∅}|; the overlap volume |V|; the fraction of the tumor it occupies, |V| / |T|; the distance to the nearest eloquent region, the minimum of dₛ over s ∈ E; the largest regional tumor load, the maximum of ℓₛ over s ∈ E; the summed regional tumor load, the sum of ℓₛ over s ∈ E; and an ordinal Sawaya grade [1] equal to 3 when |V| > 0, 2 when the nearest eloquent region lies within 10 voxels, and 1 otherwise.

##### Laterality and Midline Features

Nine patient-level features characterized tumor laterality and midline involvement. These features quantified the hemispheric distribution of tumor and its relationship to the midline in Haskins template space. Detailed definitions of the nine laterality and midline features are given below.

Each Haskins region carries a left, right, or midline hemisphere label, and fₗ, fᵣ, and fₘ denote the fractions of |T| lying in the regions of each. The nine features are fₗ, fᵣ, and fₘ; the laterality index (fₗ − fᵣ) / (fₗ + fᵣ), set to zero when both vanish; midline crossing, equal to one when fₗ > 0.05 and fᵣ > 0.05; corpus callosum involvement, equal to one when the tumor intersects any corpus callosum label; the corpus callosum fraction of the tumor; the signed displacement of the tumor centroid from the brain centroid along the left–right axis, in voxels; and the number of the 14 macroregions the tumor overlaps.

##### Tumor Morphometric Features

Fourteen tumor morphometric features were calculated from the tumor core. Nine are standard shape descriptors, two describe surface irregularity, and three are the volumes of the enhancing, nonenhancing, and cystic components. The whole tumor mask used for spatial localization included the enhancing, nonenhancing, and cystic components and excluded peritumoral edema.

##### Regional Statistical Analysis

To quantify the spatial patterns observed in the probabilistic maps, regional tumor overlap was compared across age, molecular subtype, EOR, and sex at both the 107-region and 14-macroregion resolutions. Overall differences among groups were assessed using the Kruskal–Wallis test. Group-versus-rest comparisons were performed using the Mann–Whitney U test, with rank-biserial correlation reported as the effect size.

Benjamini–Hochberg correction was applied within each variable and spatial resolution. A false-discovery-rate–corrected *q* value below 0.05 was considered statistically significant.

The non-*BRAF* molecular subgroup maps were exploratory because of the smaller numbers of patients within individual molecular categories and the nonexclusive assignment of patients with co-occurring alterations.

##### Pediatric Atlas Registration and Quality Control

Images were preprocessed as follows [2]: MRI scans from each patient were oriented to the left-posterior-superior (LPS) coordinate system, the T1-weighted, T2-weighted, and T2-weighted fluid-attenuated inversion recovery (T2-FLAIR) images were aligned with the corresponding post-contrast T1-weighted image, and these images were uniformly resampled to a 1 mm³ isotropic resolution using the anatomical SRI24 atlas through the Greedy algorithm, as facilitated by the open-source Cancer Imaging Phenomics Toolkit (CaPTk, version 1.8.1). Because preprocessing had already placed every image in SRI24 space, a single rigid transform from SRI24 space to the Haskins pediatric template [3] was computed with FreeSurfer mri_robust_register [4] and applied to every patient. Each available structural MRI sequence and the template were parcellated with SynthSeg+ [5], and the sequence was deformably registered to the template with EasyReg [6,7] using those parcellations. Registration quality was assessed separately for each sequence using brain Dice, defined as the Dice overlap between the whole-brain masks derived from the registered patient and template parcellations. For each patient, the deformation field from the sequence with the highest brain Dice was retained for subsequent analyses.

Registrations were visually reviewed in ascending order of brain Dice by reviewers blinded to molecular subtype and outcome. Rather than using a prespecified cutoff, the exclusion threshold was defined at a natural gap in the brain Dice distribution, below which registrations showed anatomically unacceptable alignment. The resulting brain Dice threshold was 0.92. Among retained registrations, the median brain Dice was 0.986 (IQR, 0.984–0.987).

The selected transformation was used to map the tumor segmentation to Haskins template space using nearest-neighbor interpolation to preserve discrete label values. All probabilistic maps and atlas-derived spatial features were generated in Haskins template space.

##### Probabilistic Mapping

For each patient, the registered whole tumor mask was converted to a binary tumor mask in Haskins template space. The whole tumor mask comprised enhancing, nonenhancing, and cystic tumor components; peritumoral edema was excluded.

Registered binary tumor masks were averaged voxel-wise within each group to generate probabilistic maps. At each voxel, the resulting value represented the proportion of patients in the corresponding group whose tumor involved that location.

Probabilistic maps were generated by age, molecular subtype, EOR, and sex. Age groups were 0–2, 2–5, and 5–18 years, chosen to reflect developmental stages rather than equal intervals. Molecular groups were *KIAA1549–BRAF* fusion, *BRAF* V600E, and non-*BRAF*. EOR groups were GTR, STR, and biopsy/no-surgery. Patients without available molecular subtype were excluded from molecular subtype analyses.

Within the non-*BRAF* group, additional probabilistic maps were generated for eight molecular categories using multi-hot encoding, such that patients with more than one applicable alteration contributed to each corresponding category.

Display scaling of the probabilistic maps was applied only for visualization and did not alter the underlying tumor overlap values or the regional statistical analyses.

##### Voxel-wise t Statistic Maps

For each group, a voxel-wise Welch t statistic compared tumor occurrence with the remaining patients in the corresponding analysis set. These t statistic maps displayed the direction and magnitude of spatial differences between groups.

Statistical inference for differences in tumor location was based on the region-level analyses described above.

##### EOR Classification

The Discovery (n = 273) and Replication (n = 76) partitions were those of the previously published clinico-imaging risk stratification models [2,8]. Two XGBoost classifiers were evaluated. The primary classifier distinguished GTR from STR or biopsy/no-surgery, and the secondary classifier distinguished biopsy/no-surgery from STR among patients in those two EOR groups. Candidate predictors included atlas-derived spatial features, tumor morphometric features, and clinical variables including age, sex, and NF1 status. For each EOR comparison, correlated continuous features were grouped using Spearman rank correlation with a cutoff of |ρ| = 0.70 and complete linkage, and the feature with the largest rank-biserial correlation was retained from each correlated group. Missing values were imputed with the median for continuous features and the mode for binary features, and continuous features were then standardized, with both steps fit within the training data of each fold. Feature clustering and selection for each comparison were run within each outer training fold of the Discovery partition, so the Replication partition was used solely for final scoring.

Model performance in the Discovery partition was evaluated using stratified nested cross-validation with five outer folds repeated five times and five inner folds. The final classifier was retuned using the complete Discovery partition and evaluated once in the Replication partition. Probabilities were calibrated using Platt scaling based on out-of-fold predictions. Model performance was summarized using the area under the receiver operating characteristic curve (AUC), Brier score, and calibration slope. Shapley additive explanations (SHAP) analysis was used to assess feature contributions. The EOR classifiers characterize associations between preoperative features and the EOR recorded in this retrospective cohort and do not estimate the EOR that is safe or achievable for an individual patient.

##### Clinico-Imaging Risk Stratification

To evaluate whether the 107 atlas-derived regional overlap features could be used in place of the manually assigned tumor location variable, that variable was replaced with the features within two previously published pLGG clinico-imaging risk stratification models. The clinicoradiomic model combined clinical predictors with multiparametric radiomic features [2], and the clinico-ResNet model combined the same clinical predictors with deep learning image features [8]. For each model family, the model containing the manually assigned tumor location variable was compared with a corresponding model in which that variable alone was replaced with the 107 atlas-derived regional overlap features.

The clinical predictors, imaging features, preprocessing, patients, outcome definition, and outer cross-validation folds remained unchanged between paired comparisons. Each of the four feature sets was fit with a penalized Cox model [9], an elastic-net Cox proportional hazards model whose mixing ratio and regularization strength were selected inside each outer fold. Because the replacement introduced 107 atlas-derived regional overlap features in place of one manually assigned tumor location variable, the regional overlap feature block was assigned a separate penalty factor selected within Discovery cross-validation; the selected factor was 0.554 for the clinicoradiomic model and 0.628 for the clinico-ResNet model. The 107 features passed through the same imputation, standardization, zero-variance filtering, and correlation pruning as the imaging block, and 83 of the 107 survived that pruning and entered the model.

The Discovery partition included 273 patients with 81 progression-free survival (PFS) events, and the Replication partition included 76 patients with 31 PFS events. Model performance was evaluated using the Harrell [10] and Uno [11] concordance indices (C-indices), giving four paired comparisons: two imaging feature sets × two C-indices. Cross-validation used 10 stratified outer folds and 10 inner folds, identical across all four feature sets, so the per-fold values are paired. Uno's index was truncated at the 85th percentile of the fitting rows' event times, which for the Replication report is 49.2 months. The Discovery interval is a 95% confidence interval (CI) over the 10 outer-fold values, and the Replication interval a 500-sample percentile bootstrap over Replication patients. Paired outer-fold differences between models containing the manually assigned tumor location variable and models containing the 107 atlas-derived regional overlap features were assessed using the corrected resampled paired t test [12].

##### Hyperparameter Search Spaces

EOR classifiers. Both XGBoost classifiers were tuned using Tree-structured Parzen estimator search over 60 trials within each inner fold, maximizing the inner-fold AUC. The search space included number of estimators from 100 to 500 in increments of 50; maximum depth from 2 to 5; learning rate from 0.01 to 0.3 on a logarithmic scale; subsample fraction from 0.6 to 1.0; column subsample fraction from 0.6 to 1.0; minimum child weight from 1 to 10; L2 regularization from 10⁻³ to 10 on a logarithmic scale; L1 regularization from 10⁻³ to 10 on a logarithmic scale; and split-loss threshold from 10⁻³ to 5 on a logarithmic scale. Class imbalance was accounted for using the positive-class scale weight. After retuning on the complete Discovery partition, the primary classifier distinguishing GTR from STR or biopsy/no-surgery used 250 estimators, maximum depth 3, learning rate 0.167, subsample fraction 0.730, column subsample fraction 0.837, minimum child weight 2, L2 regularization 0.047, L1 regularization 2.113, and split-loss threshold 1.660. The secondary classifier distinguishing biopsy/no-surgery from STR used 100 estimators, maximum depth 5, learning rate 0.043, subsample fraction 0.748, column subsample fraction 0.964, minimum child weight 2, L2 regularization 2.645, L1 regularization 0.414, and split-loss threshold 0.081.

Penalized Cox models. The elastic-net mixing ratio was selected from 0.1, 0.3, 0.5, 0.7, 0.8, 0.9, and 1.0. Penalty strength was selected from a 100-point path descending to one tenth of the strength at which all coefficients were zero, with a convergence limit of 50,000 coordinate-descent iterations. Clinical predictors were assigned a penalty factor of 0.3. The 107 atlas-derived regional overlap features were assigned a separate penalty factor selected from 40 values logarithmically spaced between 0.01 and 100 by cross-validation within the Discovery partition.

Reproducibility. Random number generators were seeded at 20260705 for the EOR classification analyses and 42 for the clinico-imaging risk stratification analyses. The machine learning analyses were performed using Python 3.12.12, NumPy 2.2.6, SciPy 1.18.0, statsmodels 0.14.6, scikit-learn 1.9.0, XGBoost 3.3.0, PyTorch 2.13.0, Polars 1.43.0, lifelines 0.30.0, scikit-survival 0.28.0, Optuna 4.9.0, SHAP 0.52.0, and Matplotlib 3.11.0.

#### Supplementary Table S1. Mapping of the 107 Haskins Regions to 14 Anatomic Macroregions

The 107 Haskins regions were grouped into the following 14 prespecified anatomic macroregions:

| **Anatomic macroregion** | **FreeSurfer label (Haskins parcellation)** | **n** |
| --- | --- | --- |
| Brainstem | Brain-Stem | 1 |
| Cerebellum | L/R Cerebellum-White-Matter; L/R Cerebellum-Cortex | 4 |
| Corpus callosum | CC_Posterior; CC_Mid_Posterior; CC_Central; CC_Mid_Anterior; CC_Anterior | 5 |
| Cingulate cortex | L/R caudalanteriorcingulate; L/R isthmuscingulate; L/R posteriorcingulate; L/R rostralanteriorcingulate | 8 |
| Frontal cortex | L/R caudalmiddlefrontal; L/R lateralorbitofrontal; L/R medialorbitofrontal; L/R paracentral; L/R parsopercularis; L/R parsorbitalis; L/R parstriangularis; L/R precentral; L/R rostralmiddlefrontal; L/R superiorfrontal; L/R frontalpole | 22 |
| Insular cortex | L/R insula | 2 |
| Occipital cortex | L/R cuneus; L/R lateraloccipital; L/R lingual; L/R pericalcarine | 8 |
| Parietal cortex | L/R inferiorparietal; L/R postcentral; L/R precuneus; L/R superiorparietal; L/R supramarginal | 10 |
| Temporal cortex | L/R bankssts; L/R entorhinal; L/R fusiform; L/R inferiortemporal; L/R middletemporal; L/R parahippocampal; L/R superiortemporal; L/R temporalpole; L/R transversetemporal | 18 |
| Deep gray nuclei | L/R Thalamus-Proper; L/R Caudate; L/R Putamen; L/R Pallidum; L/R Accumbens-area; L/R VentralDC | 12 |
| Limbic and medial temporal structures | L/R Hippocampus; L/R Amygdala | 4 |
| Optic pathway and suprasellar region | Optic-Chiasm | 1 |
| Vascular structures | L/R vessel | 2 |
| Ventricular structures and CSF | L/R Lateral-Ventricle; L/R Inf-Lat-Vent; 3rd-Ventricle; 4th-Ventricle; CSF; L/R choroid-plexus; 5th-Ventricle | 10 |
| **Total** |  | **107** |

Region names are the FreeSurfer labels produced by SynthSeg+ in Haskins template space: 68 cortical parcels of the Desikan-Killiany atlas (34 per hemisphere, prefixed ctx-lh- and ctx-rh-) and 39 subcortical, ventricular, and midline labels. L/R denotes the paired left and right labels.

#### Supplementary Table S2. Atlas-Derived Spatial and Tumor Morphometric Feature Set

| **Feature family** | **No. of features** | **Definition** |
| --- | --- | --- |
| Regional tumor overlap | 107 | Proportion of each Haskins region occupied by tumor |
| Minimum tumor-to-region distance | 107 | Minimum Euclidean distance from the tumor to each Haskins region; 0 mm when tumor intersects the region |
| Macroregion tumor overlap | 14 | Tumor overlap summarized across each of the 14 anatomic macroregions |
| Macroregion minimum distance | 14 | Minimum tumor-to-region distance for each of the 14 macroregions |
| Eloquent-region summaries | 7 | Summary measures of tumor involvement across 30 Haskins regions designated as eloquent |
| Laterality and midline features | 9 | Measures characterizing hemispheric distribution and midline involvement |
| Tumor morphometric features | 14 | Measures of tumor shape and the volumes of enhancing, nonenhancing, and cystic components |
| Total | 272 | Atlas-derived spatial and tumor morphometric features |

Clinical variables used in the EOR classifier, including age, sex, and NF1 status, were analyzed separately and were not included in the 272-feature total.

#### Supplementary Table S3. Haskins Regions Designated as Eloquent

Thirty of the 107 Haskins regions were prespecified as eloquent. The seven eloquent region summary features were computed across these regions.

| **Sawaya grade III category** | **Eloquent Haskins regions** | **n** |
| --- | --- | --- |
| Motor and sensory cortex | L/R precentral; L/R postcentral; L/R paracentral | 6 |
| Speech center | L/R parsopercularis; L/R parstriangularis; L/R superiortemporal; L/R supramarginal | 8 |
| Visual center | L/R pericalcarine; L/R cuneus; L/R lingual | 6 |
| Basal ganglia | L/R Caudate; L/R Putamen; L/R Pallidum | 6 |
| Thalamus | L/R Thalamus-Proper | 2 |
| Brainstem | Brain-Stem | 1 |
| Other | Optic-Chiasm | 1 |
| **Total** |  | **30** |

These categories correspond to the grade III eloquence categories of Sawaya et al. [1], applied at the resolution of the parcellation.

#### Supplementary Analyses

##### S1. PFS by EOR

Kaplan–Meier curves compared patients with GTR with patients with STR and biopsy/no-surgery (Supplementary Figure S1). Median PFS was 147.1 months for GTR and 45.1 months for STR or biopsy/no-surgery (log-rank χ² = 56.0; *P* = 7.2 × 10⁻¹⁴).

##### S2. Sex-Stratified Probabilistic Maps

Probabilistic maps and regional analyses were generated for female (n = 156) and male (n = 193) patients (Supplementary Figure S2). Both groups showed similar predominant posterior fossa and suprasellar tumor distributions. No anatomic macroregion differed significantly between female and male patients after Benjamini–Hochberg correction (0 of 14 macroregions; smallest omnibus *q* = 0.99).

##### S3. Secondary EOR Classifier

The secondary classifier distinguished biopsy/no-surgery from STR and used 9 selected features (Supplementary Figure S3). The AUC was 0.652 (95% CI: 0.556, 0.741) on pooled Discovery out-of-fold predictions and 0.855 (95% CI: 0.717, 0.958) in the Replication partition. Sensitivity for biopsy/no-surgery was 0.431 (95% CI: 0.300, 0.561) in Discovery and 0.625 (95% CI: 0.368, 0.867) in Replication, specificity was 0.842 (95% CI: 0.759, 0.921) and 0.727 (95% CI: 0.529, 0.905), and accuracy was 0.664 (95% CI: 0.575, 0.739) and 0.684 (95% CI: 0.526, 0.842). Brier scores were 0.229 (95% CI: 0.205, 0.257) and 0.166 (95% CI: 0.123, 0.210), and calibration slopes were 0.80 (95% CI: 0.22, 1.56) and 1.92 (95% CI: 1.15, 3.71) in the Discovery and Replication partitions, respectively.

#### Supplementary Figures

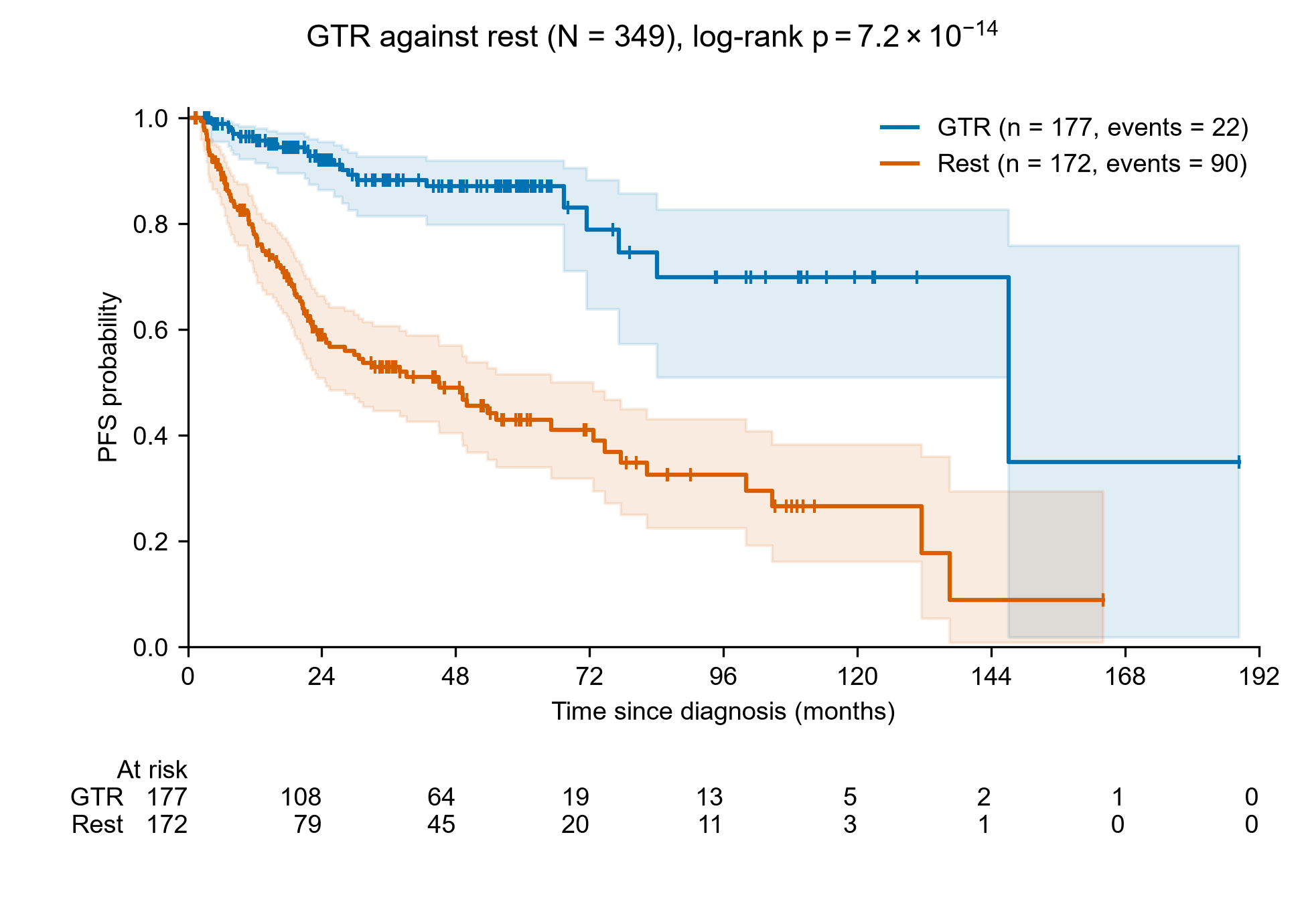

**Supplementary Figure S1. Progression-free survival by extent of resection.** Kaplan–Meier survivor functions for GTR (n = 177, 22 events) against STR or biopsy/no-surgery (n = 172, 90 events), the dichotomy the extent-of-resection classifier separates, with censoring marks, confidence bands, and numbers at risk; medians 147.1 and 45.1 months, log-rank χ² = 56.0 on 1 degree of freedom, *P* = 7.2 × 10⁻¹⁴. GTR = gross total resection/near total resection, PFS = progression-free survival, STR = subtotal resection.

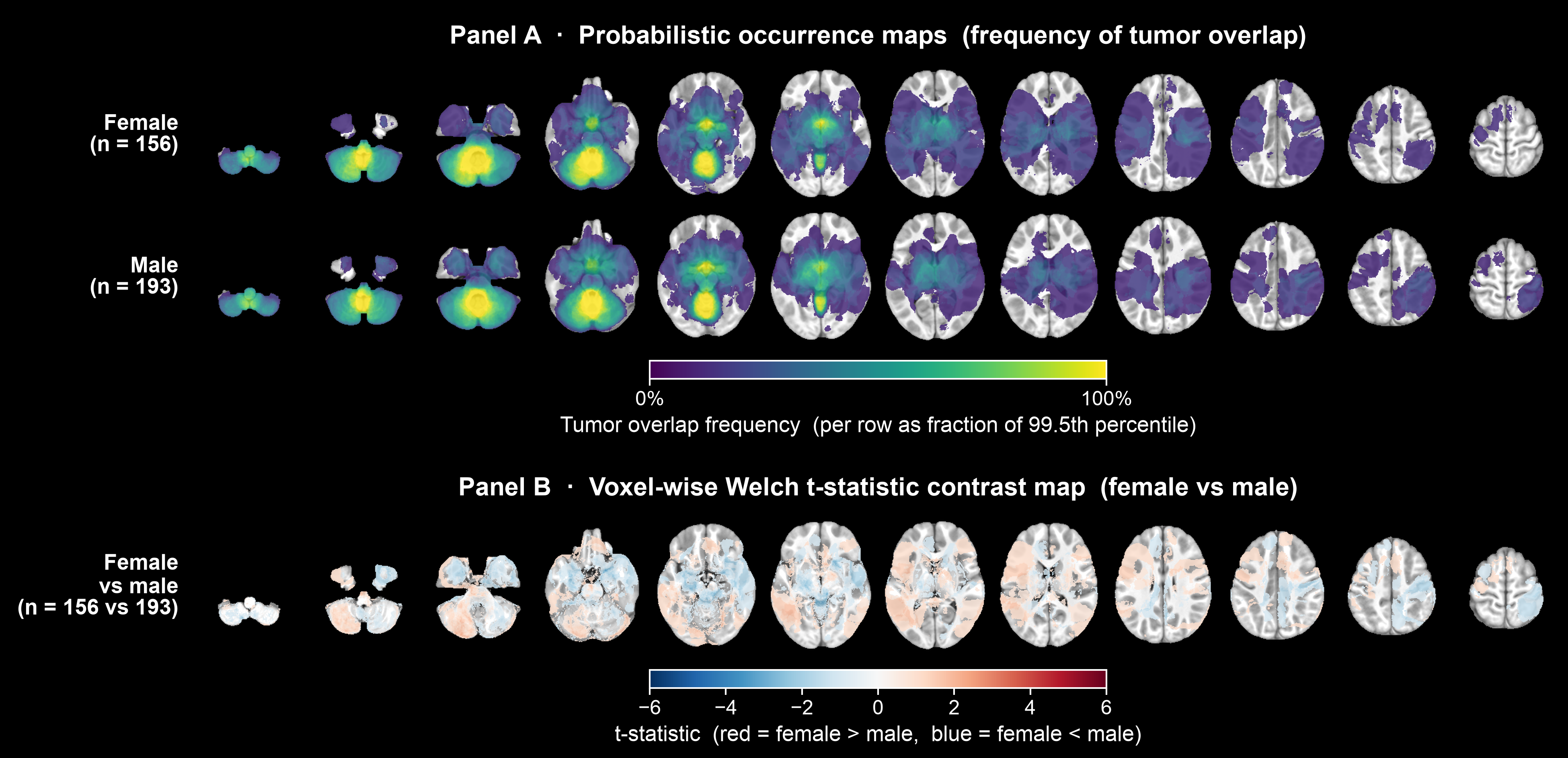

**Supplementary Figure S2. Sex-stratified maps.** (A) Probabilistic maps for female (top) and male (bottom) patients, viridis scale, scaled within each row to that row's 99.5th percentile of overlap frequency. (B) Voxel-wise Welch t statistic map comparing female (n = 156) with male (n = 193) patients, red–blue diverging scale, red indicating greater tumor probability in female patients. Both sexes show the same predominant posterior-fossa and suprasellar distribution, and no region differed between them after correction.

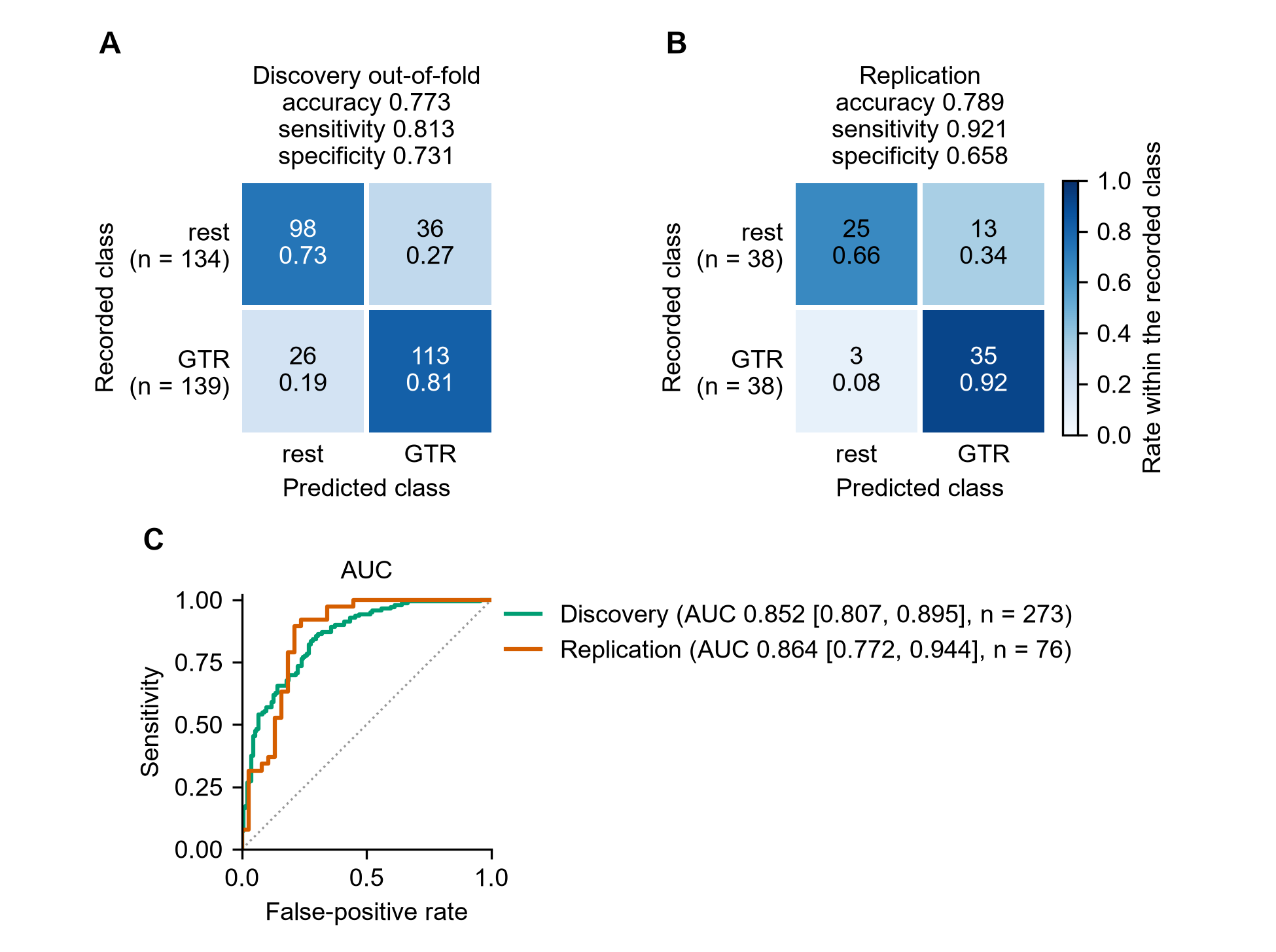

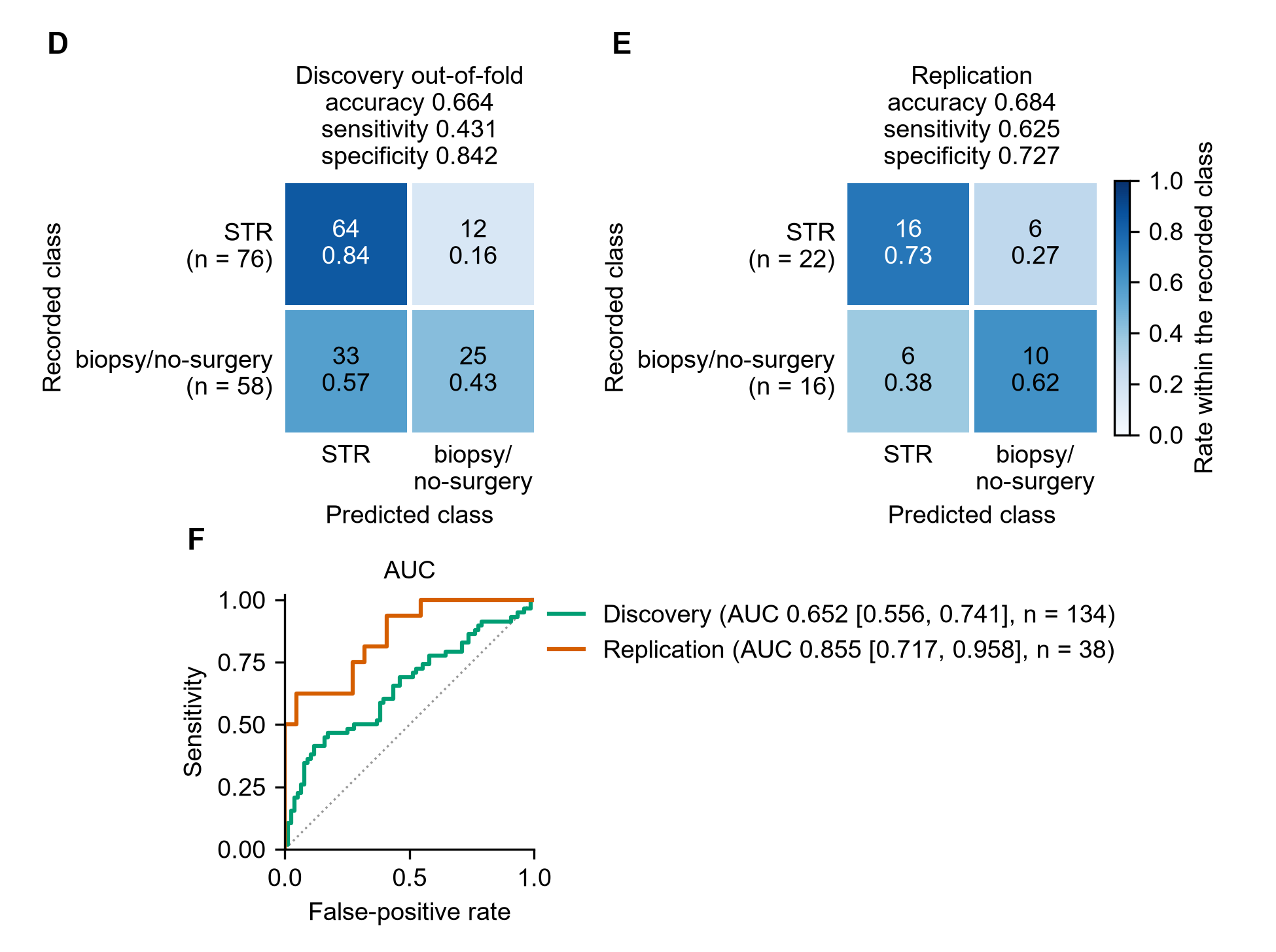

**Supplementary Figure S3. Performance of the EOR classifiers.** (A–C) Primary XGBoost classifier distinguishing GTR from STR or biopsy/no-surgery using 60 selected features. (D–F) Secondary XGBoost classifier distinguishing biopsy/no-surgery from STR using 9 selected features. For each classifier, Discovery out-of-fold (A, D) and Replication (B, E) confusion matrices and receiver operating characteristic curves (C, F) are shown. AUCs in the Discovery and Replication partitions were 0.852 and 0.864, respectively, for the primary classifier and 0.652 and 0.855, respectively, for the secondary classifier. AUC = area under the receiver operating characteristic curve, EOR = extent of resection, GTR = gross total resection/near total resection, rest = STR or biopsy/no-surgery, STR = subtotal resection.
